# Genomics in nephrology: identifying informatics opportunities to improve diagnosis of genetic kidney disorders using a human-centered design approach

**DOI:** 10.1101/2023.10.06.23296660

**Authors:** Katrina M. Romagnoli, Zachary M. Salvati, Darren K. Johnson, Heather M. Ramey, Alexander R. Chang, Marc S. Williams

**Affiliations:** Department of Population Health Sciences, Geisinger Clinic, Danville, Pennsylvania, USA; Department of Genomic Health, Geisinger, Danville, PA, USA; Departments of Nephrology and Population Health Sciences, Geisinger, Danville, PA, USA

**Keywords:** Genomic medicine, nephrology, human-centered design

## Abstract

**Background:** Genomic conditions in nephrology often have a long lag between onset of symptoms and diagnosis. To design a real time genetic diagnosis process that meets the needs of nephrologists, we need to understand the current state of the diagnostic process of genomic kidney disorders, barriers and facilitators nephrologists experience, and identify areas of opportunity for improvement and innovation.

**Methods:** Qualitative in-depth interviews were conducted with 16 nephrologists from 7 health systems across the US, with variable levels of experience with genetic testing and diagnosis. Rapid analysis identified themes in the interviews. Themes were then used to develop service blueprints (visual diagrams representing relationships between components of a service) and process maps depicting the current state of genetic diagnosis of kidney disease, helping visualize the current state, along with associated barriers and facilitators.

**Results:** Themes from the interviews included the importance of trustworthy resources, guidance on how to order tests, and evidence-based recommendations on what to do with results. Barriers included lack of knowledge, lack of access, and complexity surrounding the case and disease. Facilitators, based on current genetic testing services used by participants, included good user experience, straightforward diagnoses, and support from colleagues.

**Discussion:** The current state of diagnosis of genetic kidney diseases is suboptimal, with information gaps, complexity of genetic testing process, and complexity of disease impeding efficiency. This study highlights opportunities for improvement and innovation to address these barriers and empower clinicians who treat nephrological disease to access and use real time genetic information.

## INTRODUCTION

Genetic conditions are individually rare, but collectively common.[1] Most clinicians, particularly those not specializing in genetics, have not encountered these individually rare diseases during training. In fact, they are typically trained to prioritize more common diseases first, which may cause a genetic condition to be overlooked during the initial differential diagnosis process, leading to delays in diagnosis.[2,3] Barriers to genetic testing include insufficient experience and knowledge among nephrologists, perceived or real cost and access barriers, and lack of electronic health record (EHR) integration.[4] Our systematic review highlights the potential for clinical decision support (CDS) tools to improve the uptake of genetic services and the challenges in effectively implementing them, such as the reliance on alerts and reminders, lack of standards for genomic data integration, and underuse of implementation frameworks.[5] The review also demonstrated genetic CDS tools primarily focus on cancer care and pharmacogenomics, indicating a knowledge gap in applying genotype and family history data to other specialties, such as nephrology. Scant attention has been paid to clinician needs and workflow, which has hampered adoption of genetic diagnosis.[5] Use of implementation frameworks to objectively evaluate CDS systems in practice is uncommon, which may contribute to the poor uptake of genetic CDS tools in practice.[5,6] Understanding nephrologists’ perspectives and experience on genetic diagnosis in their clinical workflow, and how genetics should be incorporated, could enable the development of tailored CDS tools addressing the unique challenges faced by these clinicians.

Genetic testing may be helpful in cases of monogenic subtypes in a clinical category (e.g., congenital/cystic nephropathies, steroid-resistant nephrotic syndrome), positive family history, early age of onset, syndromic features, possibility of identifying a condition in which there may be targeted treatment, or as useful information to guide management or prognostication.[7] Genetic testing is also important for potential kidney donors with family history of genetic kidney disease, and to inform family planning. Even when clinical diagnosis is straightforward, as in the case of polycystic kidney disease (PKD), genetic information (such as the differences between *PKD1* truncating, *PKD2* truncating, *PKD1/PKD2* missense variants, and other genes or no mutation detected) can aid in predicting disease severity and prognosis, targeted familial testing, and treatment decision-making.[8–12] Many genetic kidney diseases, such as*COL4A3*- related autosomal dominant Alport syndrome, go undiagnosed and untreated.[13] Establishing a genetic diagnosis is important, as it enables the timely initiation of condition-specific management strategies, potentially resulting in improved outcomes.

### Objective

This project aimed to understand the current state of genetic diagnosis in nephrology. In partnership with nephrologists, medical geneticists, and informaticians, we addressed the following research questions:

1. What is the current state of diagnosis and treatment of complex genetic conditions in nephrology at multiple institutions?
2. What is the experience, from a clinician perspective, of diagnosing and treating patients with complex kidney conditions that may have a genetic cause?
3. What pain points, barriers, and facilitators exist in the process of diagnosing and treating patients with complex kidney conditions that may have a genetic cause?

## MATERIALS AND METHODS

### Design and setting

#### Human-centered design

Human-centered design (HCD), also known as user-centered design, is a collection of methodologies which include the user or recipient of a service throughout the design and implementation process.[14–16] The goal of HCD is to ensure the needs, desires, and context of the human beings, who will have to use or interact with the planned tool, resource, or service, are incorporated into the design at every stage. HCD methods were born from and are used extensively in the tech and design industries but have been increasingly used in healthcare innovation.[17–19] HCD methodologies include qualitative research to understand and empathize with the user’s current experience, use of that deep understanding of the current state to identify innovative solutions, and iterative design and testing of increasingly sophisticated prototypes with end users engaged in every stage.

#### Interview process

We used semi-structured interviews to understand the experiences of nephrologists diagnosing genetic conditions. An interview guide was developed by the study team (Supplemental Material) using an experiential phenomenological approach[20] seeking to understand the current lived experience of nephrologists diagnosing genetic kidney diseases. The interview guide was presented to the full research team for review and comment. Interview questions were informed by the literature on the barriers experienced by clinicians to conducting genetic testing. Interview topics included experience(s) with diagnosis of genetic kidney diseases and experience(s) with genetic testing in general. The interview guide included a brief demographic survey, open-ended questions, including an inquiry about what an ideal state for genetic diagnosis would look like, and prompts designed to elicit in-depth insights into the nephrologists’ experiences and perspectives regarding genetics in their clinical practice. Each interview was scheduled for 45 minutes and was conducted by a single investigator (DJ) in the presence of an experienced genetic clinician (MSW) who was available for clarification and follow-up questions.

#### Participant selection and sampling

We used a purposive sampling strategy to elicit diverse experiences and reactions to making genetic diagnoses. The population of interest in this study consisted of practicing U.S. nephrologists. We recruited nephrologists via convenience sampling at Geisinger, from non-Geisinger study team members’ organizations (University of Utah) and Twitter—a tweet inviting U.S. nephrologists using #nephtwitter. Opportunistic snowball sampling was conducted with intent to recruit participants from other institutions via the professional networks of participants. Eligible participants were contacted via email, inviting them to participate, and follow-up emails were sent to schedule the interviews.

#### Data Analysis

Episodic summary notes were created for each interview within 24 hours of interview completion. These notes captured the context and summarized the interview conversation. Study data were collected and managed in an Excel spreadsheet, using a framework based on the interview guide. Emergent themes were analyzed using content analysis and a rapid, thematic approach (RADaR: Rapid Data Analysis and Reporting)[21] by two independent reviewers (DJ and HR). Interview transcripts were coded using a template analysis approach. Summaries and exemplar quotes were entered into the study database (Excel). Themes (barriers, facilitators, and opportunities for innovation) were then summarized from the study database.

#### Human-centered design visualizations

##### Service blueprinting

Using the rapid analysis data set, the interview data were iteratively synthesized into two service blueprints representing the process of diagnosis by primary care clinicians from the perspective of nephrologists and the process of diagnosis by nephrologists.[22,23] Service blueprints are visual diagrams which represent a service being performed by mapping the relationships between the roles (such as clinicians, nurses, and patients), the tools or resources used (such as the EHR and lab tests), and the tasks performed to complete a service or task (such as ordering germline genetic testing).[23] They are helpful to illustrate the overall service design and delivery inclusive of context, as no tool or resource exists in a vacuum. They also help identify barriers to success and opportunities for improvement and innovation.

Information about roles, front-stage actions (those actions conducted within the view of the subject of the service blueprint), back-stage actions (those actions conducted out of the view of the subject of the service blueprint), and tools or resources used to support those actions were captured and summarized. This information was used iteratively to draft and revise service blueprints representing the current state of genetic diagnosis of kidney disease, which may or may not include genetic testing. To triangulate the findings, the draft maps were presented to the larger study team which included nephrologists, medical geneticists, and informaticians for their feedback, which informed updates to the maps. The maps were designed using Miro, an online visualization and collaboration tool.[24]

##### Process mapping

Workflow process mapping is another data visualization method used in HCD. It depicts the variability seen within heterogenous groups of users. Service blueprints represent the most common processes, and while they illustrate complexity and can be quite detailed, they tend to have a bird’s eye view of the larger process. Capturing the variability across different users is equally important, particularly as diagnostic processes are evolving to incorporate genetics. A workflow process map details a sequence of actions to help relevant stakeholders visualize and understand processes. Historically, process maps have been applied to health services research and quality improvement studies to help visualize those steps and pinpoint sites of intervention.[25] Using an approach adapted from Salvati et al.,[26] data about processes collected in interviews were illustrated using workflow process maps, which were presented to the study team and used to verify pathway validity.

## RESULTS

Sixteen nephrologists from seven different healthcare systems were interviewed (Geisinger, Columbia, Hattiesburg Clinic, University of Cincinnati, University of Utah, Johns Hopkins Medicine, and Georgetown University Hospital). Demographic information is included in Table 1.

**Table 1:**
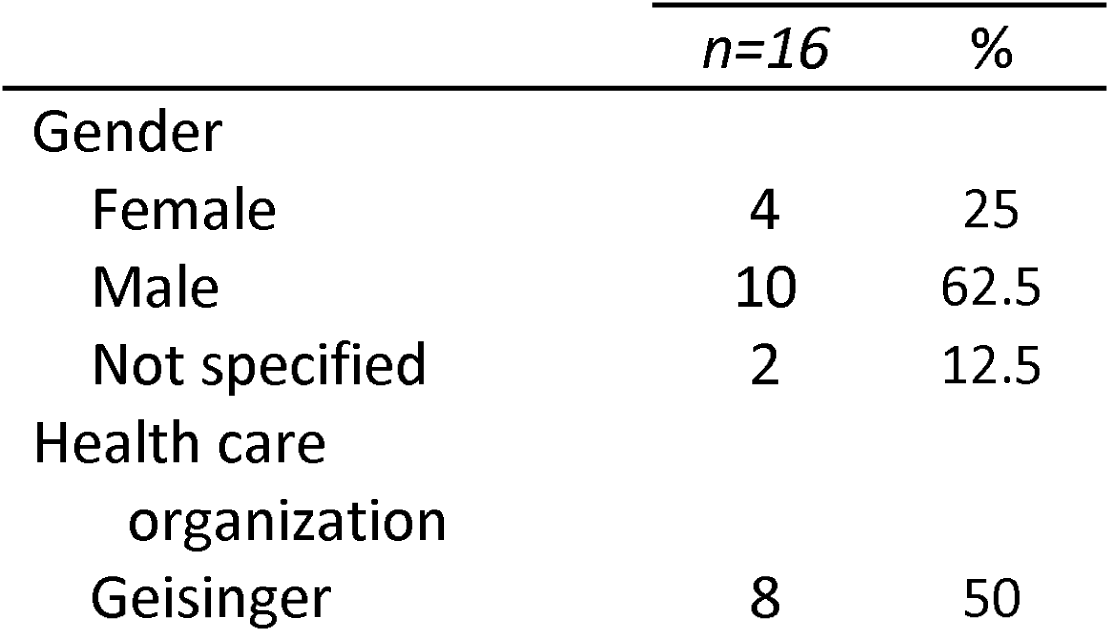
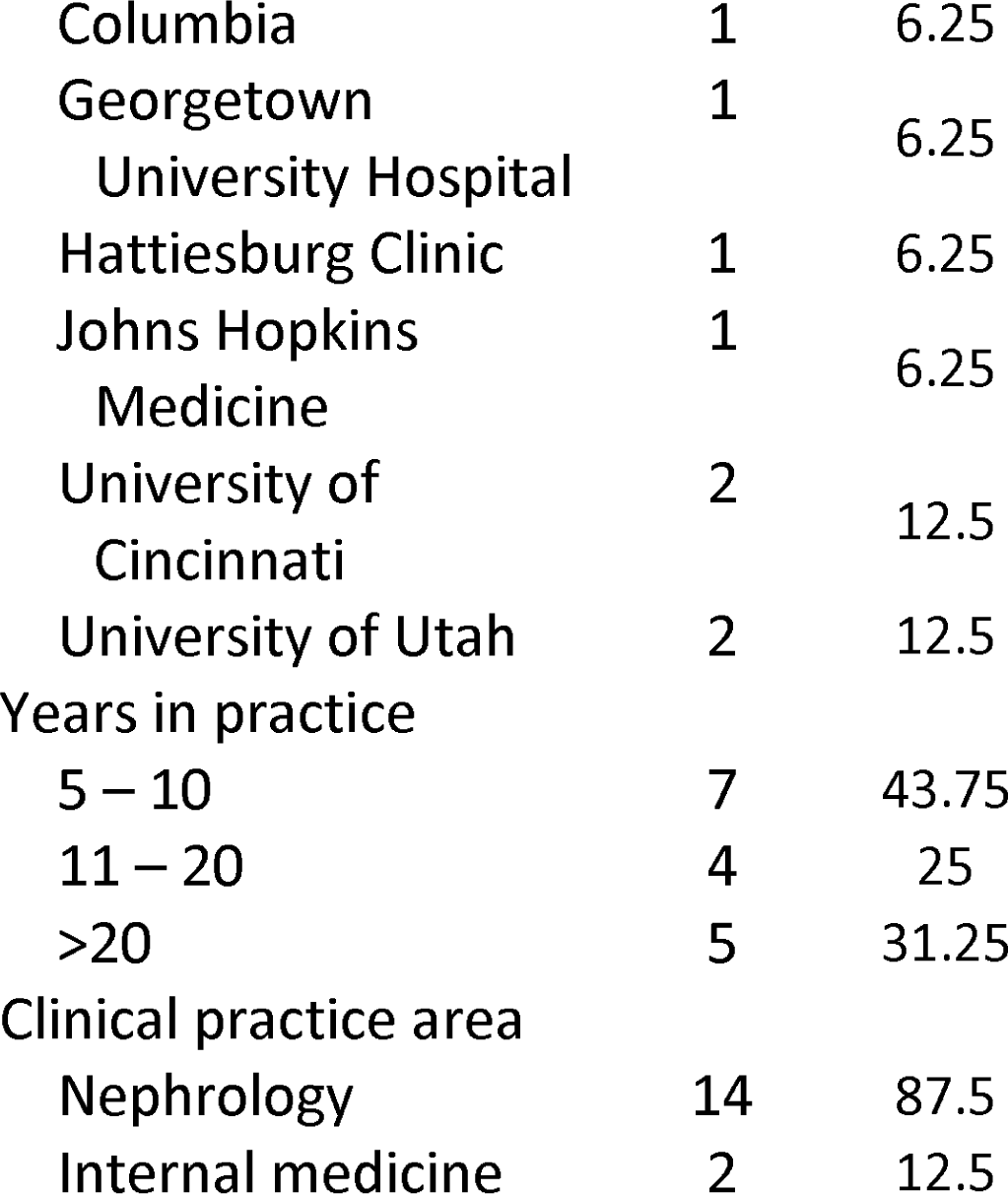
Participant demographics.

All completed the full interview. Findings include barriers to, and facilitators required for genetic diagnosis in nephrology, genetic diagnosis information needs experienced by nephrologists, and the current state of genetic diagnosis experienced by nephrologists, depicted in complementary HCD visualizations (service blueprints and workflow process maps).

### Interview themes

Themes from the interviews along with exemplar quotes from participants are summarized in Tables 2-4.

**Table 2:** Themes from interviews.

| Theme | Description | Quote |
| --- | --- | --- |
| Trustworthy genetic resources | Access to reliable genetic facilities, expert colleagues, collaborative team approaches, and vetted electronic alerts are crucial for trustworthy genetic resources | “We are fortunate enough to have an amazing facility where it’s so easy to get genetic analysis”<br>“My colleagues are probably the biggest thing...I look it up online but there’s not a ton of stuff about genetics” |
|  |  | “I would like to have more of a team approach with a genetic counselor... so they know and contribute about the workup and management of patients with genetic conditions”<br>“We have these best practice alerts which pop up in [EHR]. They can be kind of annoying, I’m not sure I would trust that type of alert unless it is really really well vetted. What else could you do? ... you can have someone still coming to see you based off the electronic algorithm saying “it looks like this person might be at risk for genetic kidney disease and somebody should consider this patient for genetic testing”. Like somebody giving them a heads up, but in a non-automatic form” |
| How to order tests | Clear guidelines in EHR, indications and patient communication aid in streamlining genetic test orders | “I use ... a renal panel which is basically in the [EHR] system... They send me a report with everything written, all references and everything I need”<br>“You would have something that could kind of tell you that this seems like a possible indication to order a genetic test and then it would have some way to inform the clinician on what [they] should talk to the patient about and how to order the genetic test and |
|  |  | what type of genetic test would be important” |
| Next steps after receiving results | Coherent reporting and referrals to genetic counselors are crucial for understanding post-test steps | “With [Genetic Counselors] I know I’m going to get a coherent nice report.”<br>“I think it’s very important for the patient to go to genetic counseling, I would refer them to genetic counselors” |

**Table 3:** Barriers to integrating genetics in clinical practice.

| Theme | Description | Quote |
| --- | --- | --- |
| Limited understanding of genetics and application in clinical care | Inadequate training and assumptions in genetics, alongside patient misunderstanding, hinder genetics integration in clinical practice | “For me, ordering genetic testing on my patients ... is not something that I learned in my training in nephrology fellowship.”<br>“People that have genetic conditions that affect the kidneys ... have usually been diagnosed before I see them”<br>“A lot of people don’t want the testing because they don’t understand the implications which is why I refer them to genetics. They’re more trained to have that conversation”<br>“I don’t think [patients] get any results until I get the results first ... are we missing any patients and they don’t actually know what the results are?” |
| Difficulty accessing genetic testing resources | Challenges in ordering, billing and limited genetic specialist | “It’s not streamlined. Every time we’re considering a |
|  | access hinder genetic testing integration | diagnosis there is running down the hall and trying to ask, "how do you guys order this?" Should I send them to genetics, should I order on my own? If I just order this in the computer am I going to get a result, is [the patient] going to get a big bill because I ordered this wrong?"<br>"I don't know how to order [genetic testing]...very often they don't send the right sample or it's not even a genetic test, it's an enzyme for a genetic test ... I'm always worried if I order it, usually not available in [EHR] but if it is I'm worried it's not going to be sent correctly to me or the patient."<br>"We lack the ability to reach out to geneticists or genetic counselors, they are extremely rare" |
| Difficulty understanding individual cases and diseases | Navigating inconclusive results, premature conclusions, and handling multi-faceted complex cases challenge clinical genetics integration | "... but still things don't add up, I don't know a diagnosis for them"<br>"Physicians I've worked with did genetic testing and they would get some results back that it's a variant of unknown significance and oftentimes I noticed they would pretty much just jump to the conclusion that's the cause even if the evidence is not quite there"<br>"I would say all the cases are diagnostically complex, |
|  |  | because by te time they come to [institution] they have usually already seen multiple providers...so they often come with a lot of data, imagine, urine studies, blood serology, histopathology...I would say all of them are complex” |

**Table 4:**
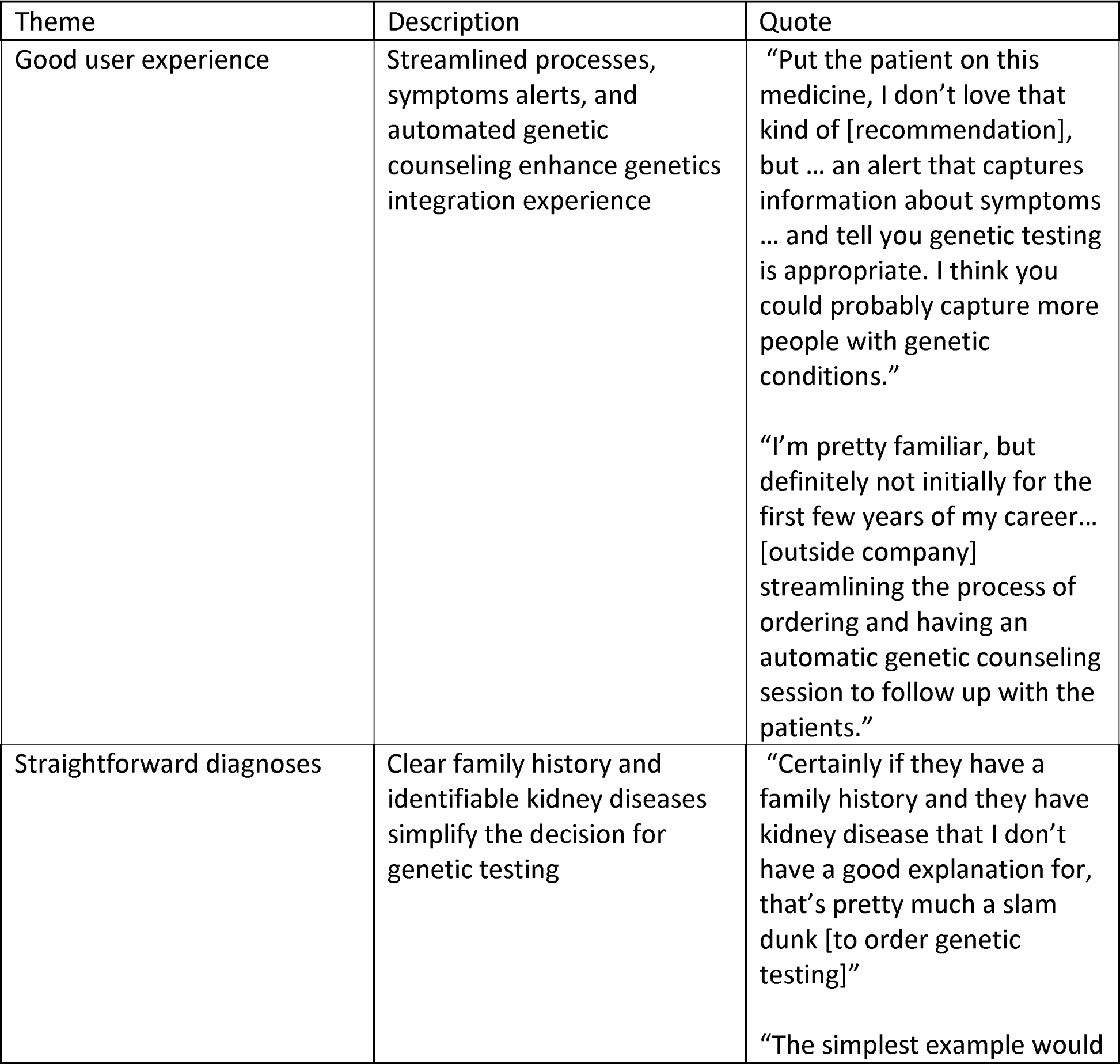
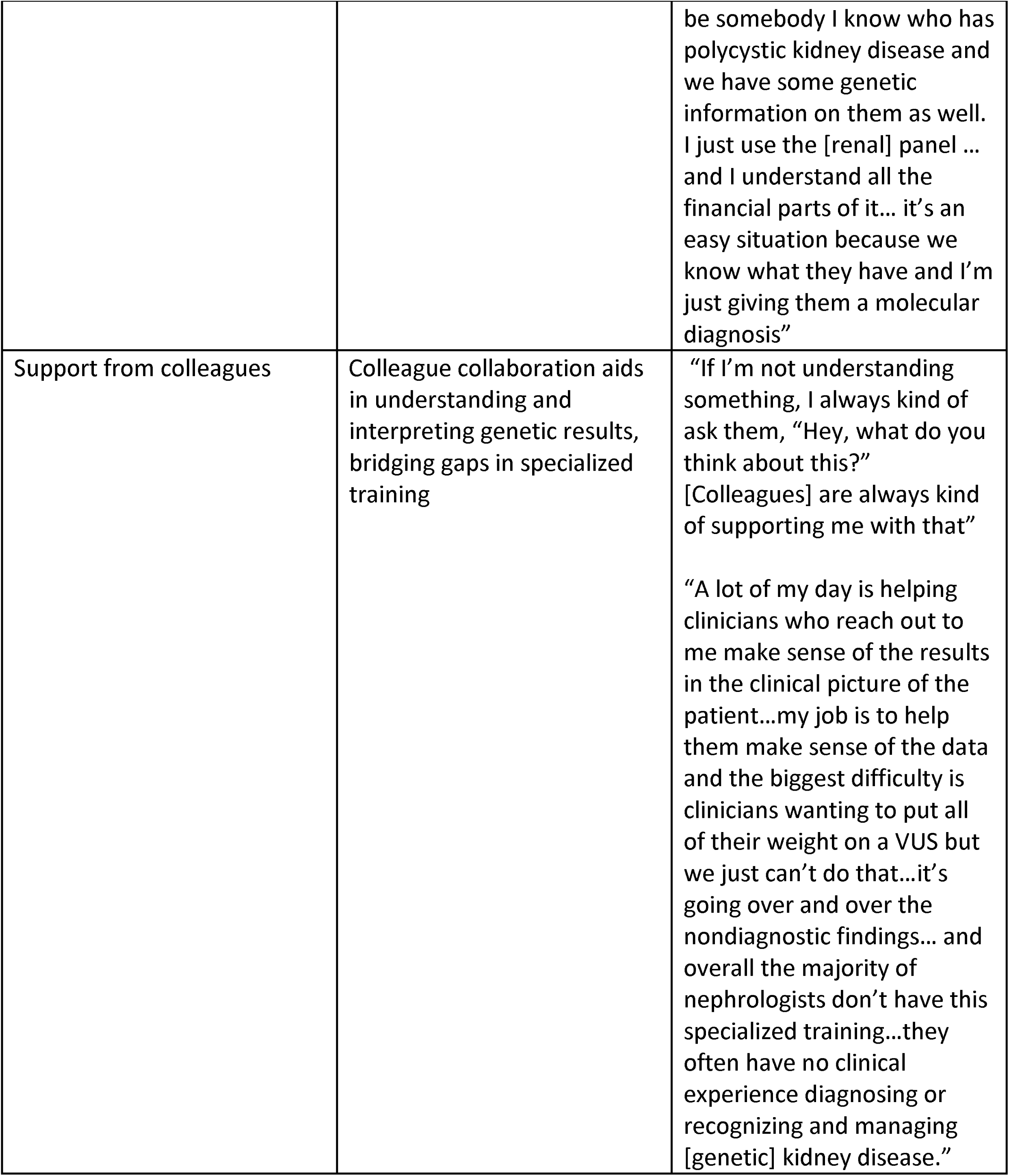
Facilitators to integrating genetics in clinical practice.

The barriers to genetic diagnosis fell into three main categories: *lack of knowledge*, *lack of access*, and *complexity surrounding the case and disease*. Nephrologists expressed challenges related to their understanding of genetics and its application in clinical care, as well as difficulties accessing genetic testing resources and grappling with the intricacies of individual cases and diseases. Facilitators required for genetic diagnosis were identified in three areas: *good user experience*, *straightforward diagnoses*, and *support from colleagues*. These factors contributed to a more seamless integration of genetics in their clinical practice.

Addressing genetic diagnosis information needs, nephrologists emphasized the importance of trustworthy resources, guidance on how to order tests, and clarity about what to do with results. The ideal future state of genetic diagnosis, as envisioned by the interviewed nephrologists, would feature a decision support tool which simplifies the genetic testing process and provides guidance on result interpretation. Additionally, many participants expressed the need for a team-based approach to genetic diagnosis, with interdisciplinary collaboration and improved knowledge surrounding genetics as key components.

### Service blueprints

While the left side of Figure 1, depicting usual practice, shows a streamlined process with minimal confusion or barriers, the right side (visualized in more detail in Figure 2), depicting the genetic diagnosis portion of the journey, is rife with complexity and barriers.

**Figure 1:**
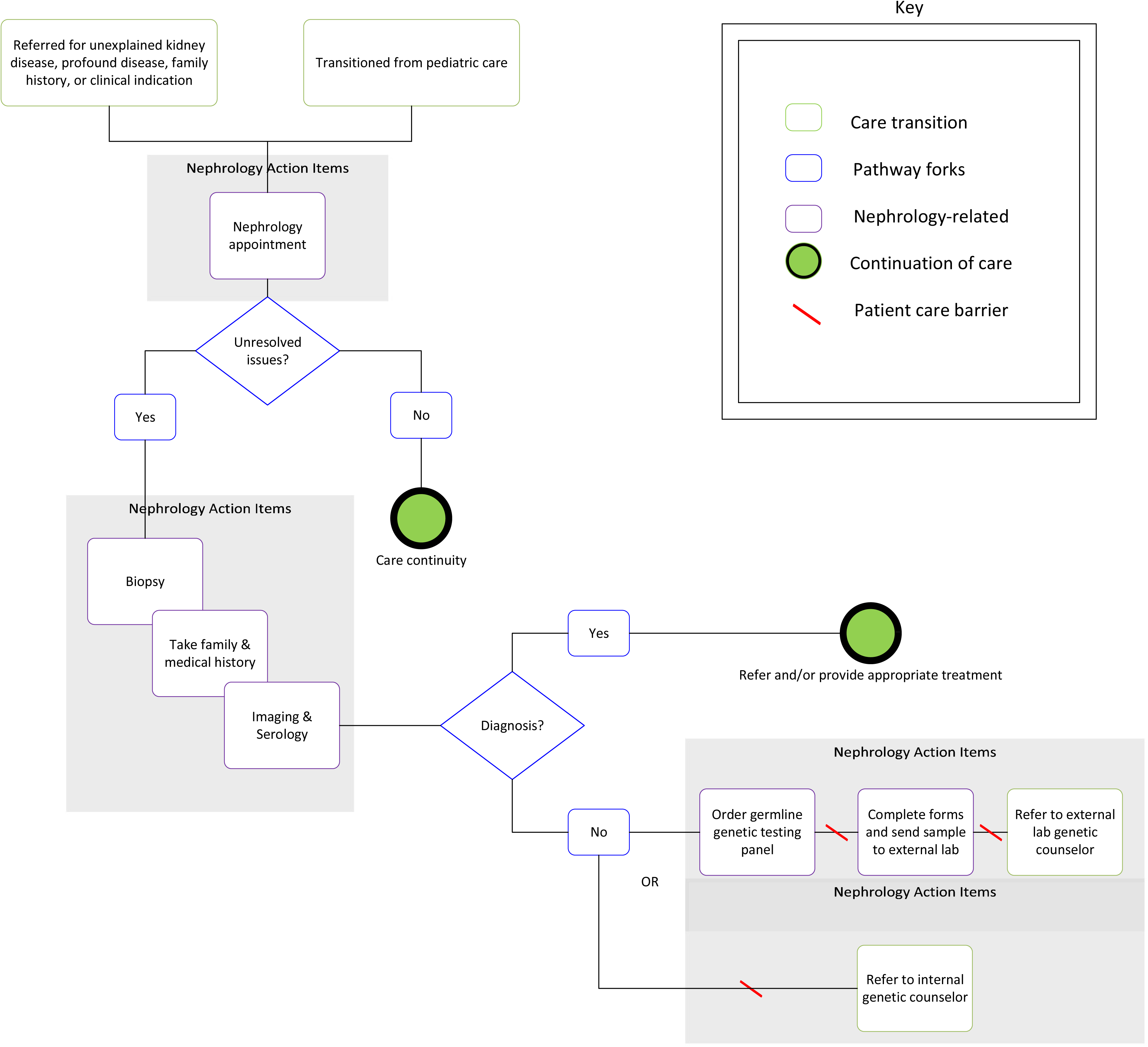
Current state of diagnosis of kidney disease by nephrologists.

**Figure 2:**
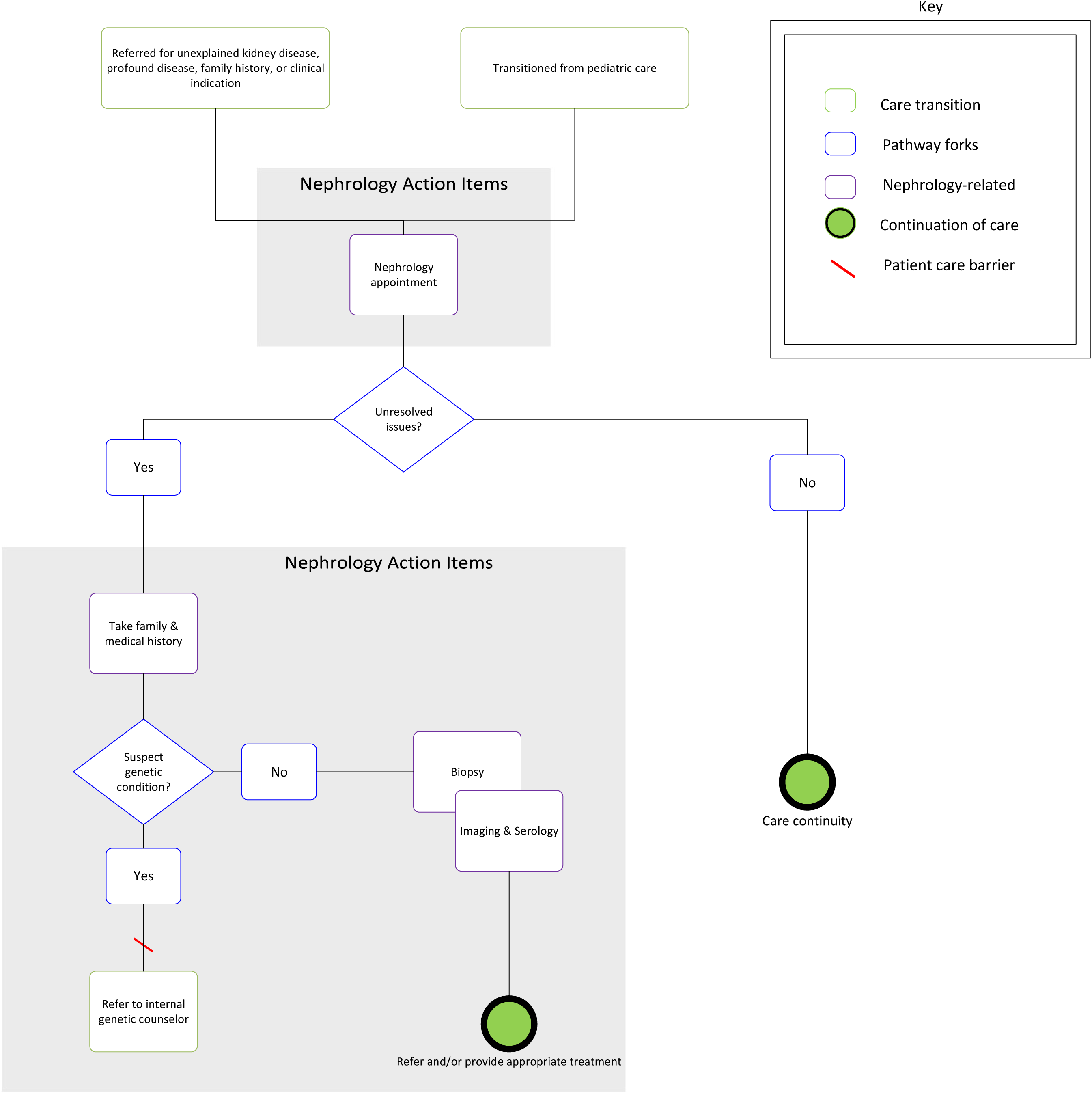
Detailed/zoomed in on genetic diagnosis section.

Service blueprints represent individual responsibilities of clinicians and personnel support on a generalized process overview, whereas workflow process maps were created to understand variability of the genetic testing process.

### Workflow process maps

Concurrent to service-blueprinting, using data from the rapid analysis following the interviews with nephrologists from seven health care systems, ZS listed process and contextual differences for ordering germline genetic testing and/or appropriately referring to a genetic counselor. These data were then iteratively adapted to workflow process maps, representing each pathway a clinician may take to diagnose genetic kidney disease. These workflow process maps were then presented to the study team of content experts, both from within and outside the health care organizations, to communicate the current state, verify pathway validity, and update maps accordingly. Ten workflow process maps were created to represent three primary processes of the current state, using nephrologists’ perspectives of ordering genetic testing across seven different health care organizations. Two organizations were found to have multiple processes nephrologists used to identify and care for patients with genetic conditions. However, it was noted from provider-stakeholder interviews these were not formal processes; rather, these steps were stakeholder-dependent due to a lack of protocol centralization. Three general workflows were identified for the current state of identifying and caring for patients with suspected genetic conditions, found from stakeholder interviews: a) Prioritizing diagnosis first, then genetic testing; b) Referral to a genetic counselor; and c) Inconsistent referrals for genetic testing (Figure 3).

All three workflow process maps showed similar approaches to why a nephrologist might order or refer a patient to receive genetic testing and how a patient who receives said testing typically presents in clinic. Genetic testing was thought to be an option for suspected rare conditions, syndromic phenotypes, genotyping polycystic kidney disease to inform prognosis, unexplained kidney disease, or in specific situations such as establishing a cause of kidney disease in potential kidney transplant recipients with CKD of unknown cause and for risk assessment of potential kidney donors. These patients will often be referred to Adult Nephrology for evaluation of CKD of unknown cause or when transitioning from pediatric to adult care.

The workflows deviate at the point where clinicians order genetic testing. One workflow is described by nephrologists prioritizing a clinical diagnosis, then ordering genetic testing if necessary (Figure 3a). This can be characterized by taking a complete medical history and family history, conducting a biopsy, imaging procedures, and/or serology. Subsequent genetic testing is warranted if a clinical diagnosis has not been made, but one nephrologist described a patient care barrier involving the return of results:

> “I don’t think [patients] get any results until I get the results first… are we missing any patients and they don’t actually know what the results are? I think ideally the provider should get the results, yet the patient should get the results in as interpretable way as possible.” As noted in the 21^st^ Century CURES Act, clinicians are required to electronically release test results to patients immediately, but this provider stakeholder mentioned patients may have a difficult time interpreting test results without provider intervention.

**Figure 3.**
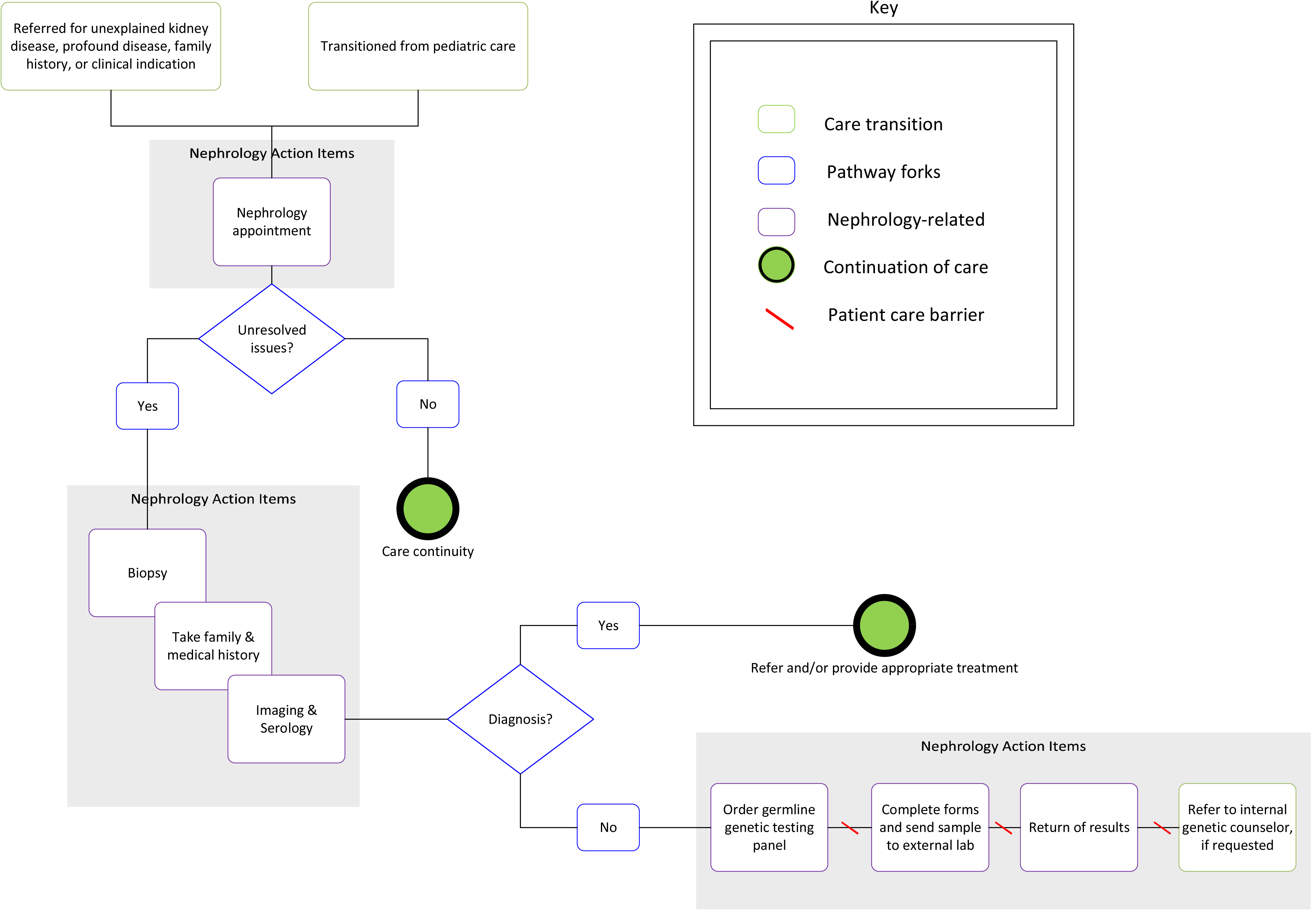
Current state workflow process map prioritizing diagnosis then genetic testing.

The second primary workflow process map shows taking the medical and family history first and referring to a genetic counselor if the nephrologist suspects a genetic condition (Figure 3b). If no genetic condition is suspected, then the typical workup of biopsy, imaging procedures, and/or serology are conducted to identify a clinical diagnosis. Patient care barriers were identified related to ordering genetic testing,

> “The process is not streamlined. There is a person running down the hall or a [Microsoft] Teams message to ask ‘how do I order this?’”, and transitioning to Genetics, “[Genetics] requires that patients fill out a patient intake form before they schedule a meeting with them. That’s a big barrier because a lot of my patients never fill them out. I feel like they should have them do it when they come [to Genetics].”

**Figure 3b.**
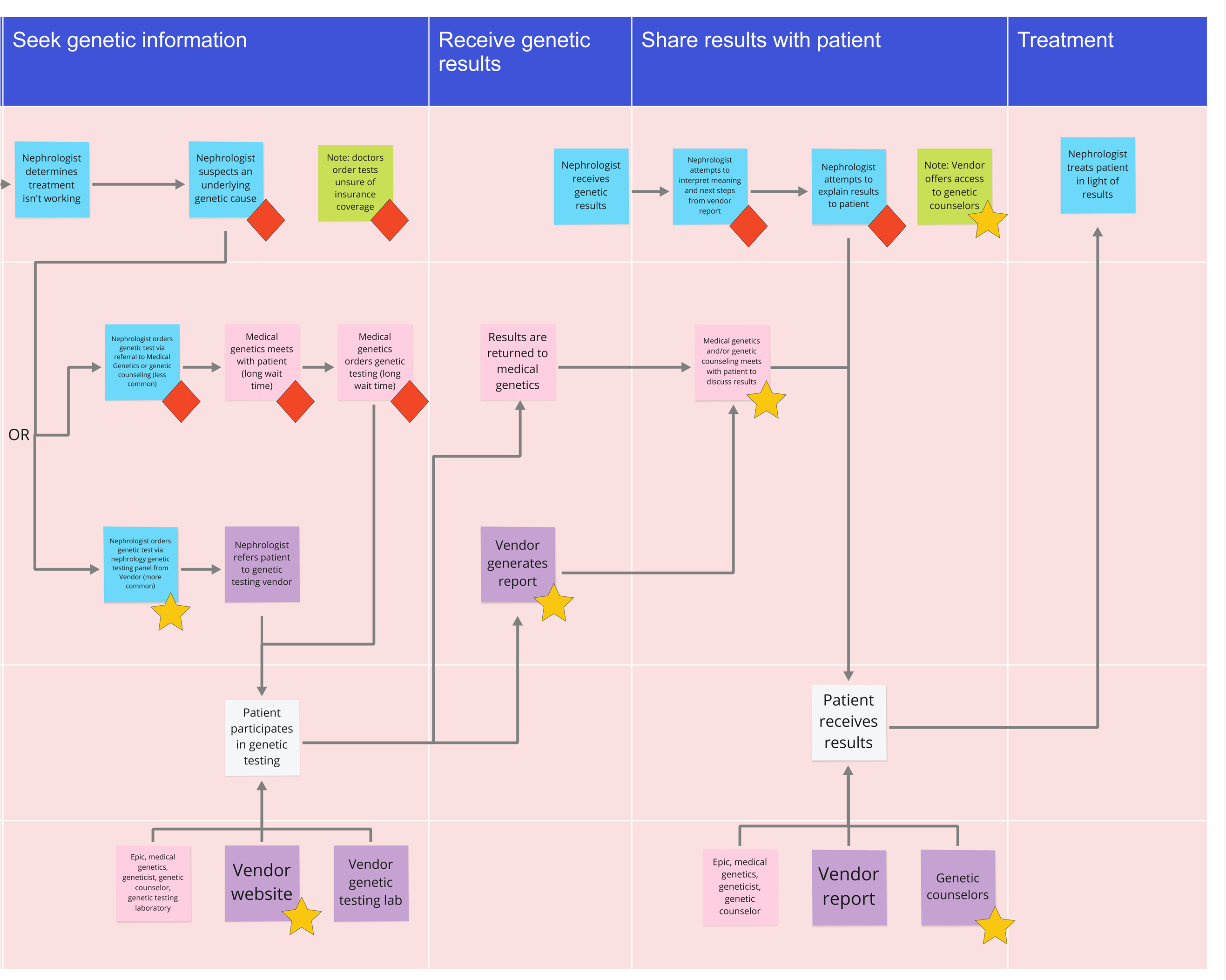
Current state workflow process map referring to a genetic counselor.

The last primary process involved ordering genetic testing themselves or referring to genetic counselors internally (Figure 3c). However, this process was variable as explained by a nephrologist from Organization 1.

**Figure 3c.**
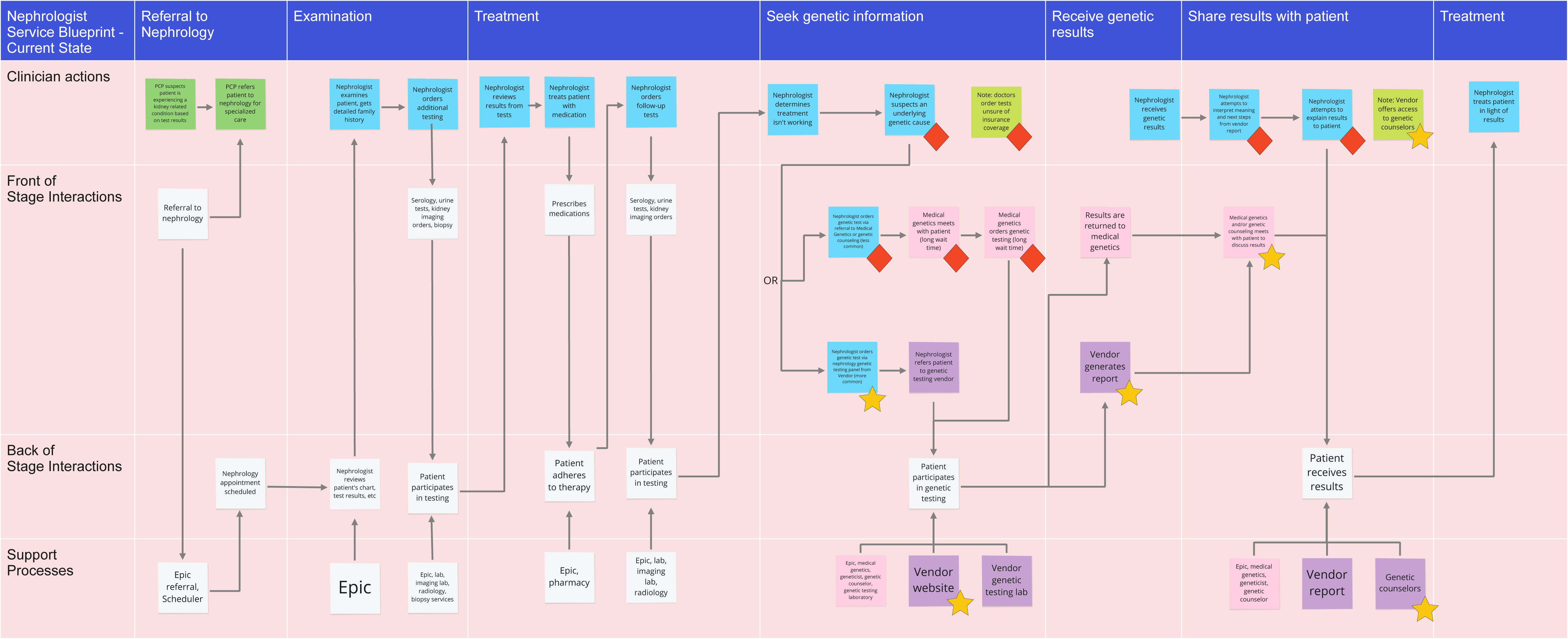
Current state workflow process map with inconsistent referrals for genetic testing.

Multiple nephrologists endorsed referring to genetic counselors internally, deferring responsibility of ordering and interpreting genetic test results:

> “In a way it’s easier to send people to Genetics because then I don’t do the testing. They do testing [and] they decide the panel they’re going to test. And I think they’re doing a really good job… they are able to disclose the information and they’re counseling the patients, right? So I kind of like that.”

Nevertheless, this provider spoke on some of the benefits to ordering genetic testing directly without referring to a genetic counselor first:

> “See, if I send the patient to Genetics…not being the one actually ordering the panel…I don’t feel like I’m learning that information, right? Whereas with [ordering directly] I actually go into their website, and I figure out, ‘OK, so these are the genes that are associated with nephrolithiasis. Interesting.’”

Overall, these descriptive, qualitative data offered varied perspectives on ordering genetic testing, or deferring that responsibility to genetic counselors, within a single organization (Organization 1). Twelve of the 16 nephrologists interviewed had experience ordering genetic testing, whereas the other four felt more comfortable referring to genetic counselors for this task. Organization 2 had two workflow process maps and Organizations 3 through 7 all had a single map representing each site. These workflows were slight variants of the three primary processes which can be viewed in the supplementary materials.

## DISCUSSION

### Summary of Findings

To understand the current state of genetic diagnosis of complex conditions in nephrology, we conducted qualitative interviews with nephrologists from 7 U.S. health systems. We identified barriers (lack of knowledge, lack of access, and complexity surrounding the case and disease) and facilitators (good user experience, straightforward diagnoses, and support from colleagues)

to timely diagnosis of genetic conditions in nephrology. Similar barriers have been identified by other research exploring why genetic testing has been poorly implemented. To identify areas of opportunity to improve genetic diagnosis, we created a suite of visual artifacts depicting the current state of genetic diagnosis of kidney conditions by nephrologists, which capture both the interaction with the larger context of healthcare and the overall service design of genetic diagnosis (service blueprints) as well as the variability of processes across different nephrologists and healthcare systems (process maps).

In an area where no guidelines or standard practices exist, we expected to see substantial variability in the steps clinicians take and this was confirmed. A systematic review of clinicians’ genetic testing practices found most studies focused on clinicians’ knowledge, attitudes, or beliefs about genetic testing, but none evaluated the experience or process of obtaining or receiving a genetic diagnosis.[27] Within nephrology, recent work reviewed the current state of evidence for the genetic diagnosis of diseases, summarizing the indications for pursuing genetic testing, and encouraging the use of genetic diagnosis in nephrology.[28] However, these do not include information on the current experience of clinicians in conducting genetic testing within any clinical area, nor specifically nephrology.

### Implications

As recognition of the role genetics has in kidney disease continues to grow, improvements to equitable access to genetic testing in nephrology practice are necessary.[7] Opportunities exist, identified in this study, to improve the process and experience of using next-generation genetic testing technology to aid in the diagnosis of genetic kidney diseases by nephrologists. These could include, but are not limited to, tools and resources which connect genetic information to clinical presentation using electronic health records at the point of care, such as provider alerts for patients who meet criteria for genetic testing, order sets to help select the most appropriate test panel for a given patient, electronic referrals to genetic counseling, and support for interpretation of genetic test results.

Yet, substantial barriers exist to the implementation of such tools. Some external testing vendors have developed nephrology-specific genetic testing products, marketing them to nephrologists directly.[29] These services include support for ordering and result interpretation, addressing some of the barriers identified in this work. However, external stand-alone services do not address other important aspects of real time genetic diagnosis design identified by participants, such as support from colleagues and management of complexity in individual cases. The large number of genes on kidney disease gene panels, while intending to be helpful by providing more information, could be difficult for clinicians to interpret due to receiving results which seem to be unrelated or having unknown significance to the indication for testing. Furthermore, genetic testing results from external services must be manually added to the patients’ health record, limiting the ability to use informational resources in the EHR that could be triggered by structured data in reports. An informatics resource leveraging genetic and phenotypic information is insufficient; nephrologists also need support in the process of deciding when and how to use genetic information, efficient and affordable acquisition of that information, and interpretation of any genetic findings, as well as the interpretation of inconclusive or absent findings.

### Limitations

This study had limitations related to sampling and data collection. First, data collection was limited to clinician stakeholders who were interested enough in genetic testing to participate in this research study from seven healthcare organizations, and participant sampling ranged from one to six stakeholder perspectives representing each site. Ultimately, a cross-case comparison by site would not have achieved thematic saturation. Rather, thematic saturation was achieved by looking at all stakeholder perspectives together, with Organization 1 representing the three primary processes for ordering germline genetic testing in Nephrology. Future studies are needed to further characterize processes across multiple institutions to ensure process saturation.

### Future Work

Future work includes conducting a design thinking workshop with nephrologists, medical geneticists, informaticians and other experts. The workshop will use the findings from the qualitative work, including the data visualizations, to build empathy and shared understanding of the current state of genetic diagnosis in nephrology among the participants. The output of the workshop will be a first draft prototype of the future state of genetic diagnosis in nephrology, using real-time genetic diagnosis innovations.

## Conclusion

The current state of genetic diagnosis in nephrology is suboptimal for timely diagnosis of genetic kidney diseases. We have identified opportunities to improve and innovate this experience with the human-centered design of a real-time genetic diagnosis tool.

## Data Availability

All data produced in the present study are available upon reasonable request to the authors

